# Large-scale neural slowing measured with electroencephalography indexes secondary thalamic degeneration in stroke

**DOI:** 10.64898/2026.06.29.26356697

**Authors:** P. R. Johnston, A. Schwarz, J. Dodd, J. D. Griffiths, J. A. Meltzer, S. C. Cramer, A. R. McIntosh

## Abstract

A central priority in stroke recovery research is the development of useful biomarkers that can reveal underlying disease states to aid diagnosis, prognosis, stratification, and treatment, ideally using tools that are readily available in clinical settings. Large-scale neural slowing has the potential to be such a biomarker, but its usefulness is currently limited by uncertainty about its nature and underlying causes. In this work, we sought to address these gaps by parameterizing abnormal resting state electroencephalography (EEG) spectral features across the scalp, and investigating their relationship to thalamic atrophy and dysfunction, using structural magnetic resonance imaging (MRI) and a computational model of corticothalamic circuit dynamics, respectively. As predicted, stroke patients (n=25) exhibited widespread spectral abnormalities, including significantly increased aperiodic exponent and offset, lower alpha frequency, and reduced beta power, which together can account for the frequently observed shift in power towards low frequencies after stroke. Furthermore, corticothalamic models fit to power spectra across the scalp inferred broad thalamic disinhibition. Crucially, these abnormalities (except beta reductions) were strongly predicted by ipsilesional thalamic atrophy measured with MRI, despite the lack of direct thalamus damage in this sample. Together, these findings highlight secondary thalamic injury as an important consequence of stroke and potential cause of widespread neural dysfunction, helping to clarify the underlying causes of post-stroke neural slowing, and demonstrating its feasibility as a clinically-accessible biomarker.

## Introduction

Developing useful biomarkers is a crucial step towards personalized medicine approaches to stroke recovery, and therefore towards reduced global burden of disability due to stroke. These biomarkers should be indicators of underlying disease states that are useful for parsing heterogeneity in research samples, and guiding clinical diagnosis, prognosis, stratification, and ultimately treatment (Boyd et al., 2017; Cramer et al., 2017). A marked shift in large-scale neural activity towards lower frequencies, detectable with electroencephalography (EEG) and magnetoencephalography (MEG), has been observed and linked to poorer patient outcomes in stroke for decades (e.g. Ahmed, 1988; Cohn et al., 1948; Johnston et al., 2023; Laaksonen et al., 2013; Sainio et al., 1983; Tecchio et al., 2005), and may hold promise as a biomarker that is non-invasive and relatively easy to measure in both research and clinical settings. Yet the underlying causes of this neural slowing remain poorly understood, limiting its utility as a biomarker.

However, recent work has better characterized this neural slowing phenomenon and clarified its potential cause. Using resting state MEG recordings from a sample of patients with chronic aphasia, we demonstrated that this frequently-observed shift towards lower frequencies is not a unitary phenomenon, but is best explained as a steepening of the aperiodic background, in conjunction with alpha oscillation slowing and reduction in beta oscillation power (Johnston et al., 2023). By applying neurophysiological modelling of a thalamocortical circuit to the same dataset, along with multimodal imaging of the thalamus, we found evidence that this widespread pattern of slowing is due to reduced intrathalamic inhibitory feedback related to secondary thalamic degeneration, providing a crucial link between non-invasive electrophysiology and underlying disease state (Johnston et al., 2024). Building on previous work showing that non-thalamic lesions can cause substantial degeneration of the thalamus in humans (e.g. Binkofski et al., 1996; Brodtmann et al., 2020; De Reuck et al., 1995; Hervé et al., 2005; Yamauchi et al., 2022), as well as thalamic disinhibition in rodents (Paz et al., 2010), this work demonstrated for the first time a potential role for secondary thalamus injury in causing large-scale neural slowing and post-stroke disability.

With the goal of corroborating these insights, and advancing them towards clinical utility, here we attempted to replicate and extend these findings in a different sample of stroke patients, using a more accessible neurophysiological tool (EEG), and basic structural imaging (T1w MRI). Specifically, our sample included 25 individuals with motor impairment at least 3 months post-stroke, who had either a left or right hemisphere lesion, carefully selected to exclude those with primary thalamic damage. Abnormal neural dynamics were estimated from resting state high-density EEG data and compared to 17 controls. Secondary thalamus degeneration was quantified as reduced thalamic volume compared to a normative population (Ge et al., 2024), with volumes estimated using semi-automated segmentation on T1w MRI. Building on our previous work, we quantified periodic and aperiodic neural dynamics across the scalp using *specparam* (Donoghue et al., 2020), and fit a computational model of a thalamocortical circuit to infer the underlying neurophysiology responsible for these dynamics (Abeysuriya & Robinson, 2016; Robinson et al., 2001). Our prior findings were largely supported, with stroke patients demonstrating the previously observed widespread pattern of abnormalities compared to controls, including steeper aperiodic slopes, alpha slowing, beta power reduction, and model-estimated thalamic disinhibition. Crucially, thalamus atrophy was strongly correlated with these abnormalities, despite the absence of direct thalamic damage in any of these patients. We also provide new insight into the topographical distribution of the electrophysiological abnormalities, as well as the effect of lesion location. Overall, this work demonstrates that the link between widespread electrophysiological abnormalities and secondary thalamus degeneration generalizes to a new sample with different clinical characteristics (i.e. motor impairment related to a mix of left and right hemisphere lesions), who did not exhibit direct thalamic damage. This work also demonstrates that this relationship can be detected with clinically-available tools (EEG and T1w MRI), bolstering its suitability as a biomarker for clinical use.

## Methods

### Participants

Stroke patients were drawn from a prior study (Cassidy et al., 2020), where the initial inclusion criteria were: individuals at least 18 years of age, with no contraindications to MRI, no substantial communication deficits, and no previous cranial surgery. 62 individuals with stroke and 22 controls initially met these criteria. The study protocol was approved by the University of California, Irvine Institutional Review Board, and all participants provided written informed consent. Secondary analysis of de-identified data was approved by the Research Ethics Board at Baycrest Health Sciences.

To align the patient sample to our previous work, only individuals who had a unilateral, supratentorial stroke at least three months prior to data collection were included. Three months was chosen as the cutoff, rather than the six months used previously, because this sample contained a larger proportion of patients with relatively recent strokes, and previous work suggests that the highest rate of thalamus degeneration would have already occurred within the first three months (Brodtmann et al., 2020). Since our focus was on secondary thalamic damage rather than primary damage, we excluded patients with a lesion mask that overlapped the thalamus mask, or a void with CSF-like contrast and a diameter of 4 mm or greater (Laveskog et al., 2018) inside or adjoining either thalamus. To further limit the possibility of direct thalamus damage, patients were excluded if they had an infarct elsewhere within the posterior cerebral artery (PCA) branches that supply the thalamus. To determine this, lesion masks were non-linearly registered to MNI space using ANTS (Avants et al., 2011) then compared to an atlas of vascular territories (Liu et al., 2023; level 1 atlas with sharp boundaries). Patients were excluded if the posterior or anterior thalamoperforator territory was the largest or second-largest region of overlap with the lesion. Finally, patients were excluded if their EEG data had excessive muscle artifacts that could not be removed with a combination of manual channel/epoch rejection and ICA (see “EEG collection and preprocessing” below). In total, 25 patients met the above criteria (15 males, 10 females). Their demographic, behavioural, and anatomical details are reported in Table 1.

**Table 1:** Patient characteristics and behavioural data. Ips. = ipsilesional, Contr. = contralesional.

| **Demographics** | **Mean** | **SD** |
| --- | --- | --- |
| Age | 55.1 | 16.1 |
| Time post-stroke | 1.7 y | 2.1 y |
| Sex | M:15, F:10 |  |
| Handedness | R: 23, L: 2 |  |
| Ethnicity | Asian: 3, Caucasian: 15, Hispanic: 4, Not reported: 3 |  |
| **Behavioural** | **Mean** | **SD** |
| UEFM Total | 43.8 | 12.74 |
| UEFM Proximal | 26 | 6.01 |
| UEFM Distal | 14.2 | 6.99 |
| NIHSS | 2.76 | 2.05 |
| **Anatomical** | **Mean** | **SD** |
| Lesion volume | 33,444 mm<sup>3</sup> | 41,421 mm<sup>3</sup> |
| Lesion side | R: 13, L: 12 |  |
| lps. thalamus volume | 3,250 mm<sup>3</sup> | 796 mm<sup>3</sup> |
| Contr. thalamus volume | 4,623 mm<sup>3</sup> | 857 mm<sup>3</sup> |
| lps. thalamus normative deviation | -6.75 | 1.98 |
| Contr. thalamus normative deviation | -4.3 | 1.28 |

Patients were initially matched to a sample of 22 right-handed controls, but only 17 (6 males, 11 females) were used here since 3 raw datasets could not be recovered and 2 were contaminated by intractable EEG artifacts. After all of the exclusions described above, patients and controls were comparable but not statistically matched in age (patients: mean (SD) = 55.1 (16.1), controls: mean (SD) = 66.5 (16.1), t(40) = -2.26, p = 0.03). Statistical controls for age are described in the results section where appropriate.

In addition to the neuroimaging measures described below, all patients completed the Upper Extremity Fugl-Meyer (UEFM; Fugl-Meyer et al., 1975) to assess motor impairment of the upper extremity, and the National Institutes of Health Stroke Scale (NIHSS) to assess overall neurological impairment.

### EEG collection and preprocessing

All participants completed 3 minutes of awake, eyes-open, resting-state EEG recording using a 256-electrode Hydrocel Geodesic Sensor Net (Electrical Geodesics, Inc.). Participants sat upright while fixating on a white cross displayed on a laptop in a dimmed room. They were instructed to minimize movements and speaking, and were monitored by an investigator during recording. Signals were acquired at a sampling rate of 1000 Hz using a high impedance Net Amp 300 amplifier and Net Station 4.5.3 (Electrical Geodesics Inc.).

EEG preprocessing was carried out with MNE-Python (v1.2.2, Gramfort et al., 2013; Larson et al., 2022). Continuous recordings were segmented into non-overlapping epochs of 4 s in length, then linearly detrended. Bad channels and epochs contaminated by movement artifacts were manually labelled and removed from all subsequent processing. The remaining epochs were submitted to independent components analysis (ICA; FastICA algorithm; Hyvärinen & Oja, 2000) for artifact rejection. More specifically, ICA was computed on a parallel copy of the data that was bandpass filtered between 0.5-40 Hz prior to epoching and detrending. Due to the large number of channels relative to the recording length, only the PCA components from the pre-whitening step that cumulatively explained 99% of the variance were passed to the ICA algorithm. ICA component scalp topographies, power spectra, and time series were inspected, and those matching ocular, cardiac, or muscle artifacts were rejected from the original (not bandpass filtered) epochs. After artifact rejection, any remaining bad epochs or channels were rejected with a second round of manual inspection. An average of 32.1 epochs (SD = 6.8) were retained for patients, and 32.9 (SD=10.01) for controls. AutoReject Local (v0.4.3; Jas et al., 2017) was then applied to detect any remaining momentary artifacts limited to specific epochs and channels (e.g. electrode pops) and interpolate them, as well as interpolate any channels previously marked as bad. Finally, data were re-referenced to the average of all electrodes, and power spectral densities (PSDs) were computed for each epoch × electrode using Welch’s method (window = 1 s, overlap = 0.5 s), then averaged over all epochs.

Abnormalities in the periodic and aperiodic components of the power spectrum were quantified with *specparam* (Donoghue et al., 2020), applied to each average PSD (1-45 Hz, peak width limit = 2-8 Hz, minimum peak height = 0.1, maximum number of peaks = 4, peak threshold = 1.5, no aperiodic knee). The tallest peak in the range 5-14 Hz was considered the alpha peak (the lower cutoff was extended downwards from typical definitions to accommodate previously observed alpha slowing; Johnston et al., 2023). Similarly, the tallest peak in the range 15-30 Hz was used to quantify beta activity.

All analyses were carried out using the 180 electrodes on the scalp (or subsets thereof, where specified; see Supplementary Table S1), excluding cheek and neck electrodes. When patients had left hemisphere lesions their electrode labels were swapped across the midline so patients with left and right hemisphere lesions could be analyzed together. Subsequent analyses therefore compare ipsilesional (IL) and contralesional (CL) hemispheres rather than left and right.

### Corticothalamic modelling

As described in Johnston et al., (2024), a computational model emulating the dynamics of a corticothalamic circuit was used to estimate physiological quantities underlying abnormal neural dynamics (Supplementary Figure S1). This model produces a simulated power spectrum by emulating the dynamic interactions between four populations of neurons: excitatory (e) and inhibitory (i) cortical populations, the inhibitory thalamic reticular nucleus (r), and the excitatory thalamic relay nuclei (s).

BrainTrak software was used to fit the linearized frequency domain version of the corticothalamic model to empirical PSDs from patients and controls (Abeysuriya, 2017; Abeysuriya et al., 2015; Abeysuriya & Robinson, 2016). BrainTrak uses a Metropolis Hastings Markov Chain Monte Carlo (MCMC) algorithm to sample the joint probability distribution of the model parameters to find the set of parameters with maximum likelihood. Nine parameters were estimated in total: five “gain” parameters representing the ratio of input to output within key loops of the corticothalamic circuit (G_ee_, G_ei_, G_ese_, G_esre_, G_srs_), the inverse synaptodendritic decay time (α) and rise time (β), the thalamocortical time delay t_0_, and the amplitude of the electromyographic artifact correction (A_EMG_). These parameters were then aggregated into three additional parameters summarizing the overall gain within the three large-scale loops of the model: X (intracortical loop), Y (corticothalamic loop) and Z (intrathalamic loop). The primary parameter of interest is Z, which reflects the gain within the inhibitory feedback loop between the reticular nucleus and relay nuclei of the thalamus.

We applied BrainTrak to individually fit the average PSDs at each scalp electrode for each participant (O’Connor & Robinson, 2004). The fitting range was restricted to 1-40 Hz, with no spatial variation in model parameters and MCMC chain length of 50,000 for sampling the joint probability distribution.

### Structural imaging and thalamus volume measurement

T1-weighted structural images were acquired for patients only, with a 3D magnetization-prepared rapid gradient echo sequence (MPRAGE; repetition time = 8.1–8.5 ms, echo time = 3.7–3.9 ms, 150 slices, voxel size = 1 × 1 × 1 mm^3^) on a 3T Philips Achieva scanner. A T2-weighted fluid-attenuated inversion recovery (FLAIR) scan was also collected (repetition time = 9000–11000 ms, echo time = 120–125 ms, 31–33 slices, voxel size = 0.58 × 0.58 × 5 mm3). Binary lesion masks were manually traced on the T1w image, informed by the overlaid T2w image. Infarct core and surrounding diffuse white matter injury were included in the lesion mask. T1w images were defaced with fsl_deface (Alfaro-Almagro et al., 2018).

To quantify thalamus volume, left and right thalami segmentations were first approximated using Freesurfer (recon-all; Fischl, 2012) applied to the T1w image. Binary thalamus masks were then created by extracting the left and right thalamus from Freesurfer’s automated volumetric subcortical segmentation. These masks were manually corrected according to a published protocol (Power et al., 2015) by two raters (PJ: 13 patients, JD: 12 patients). As noted previously, any patients with detectable lesions within their thalamus masks were excluded from further analysis.

To quantify the degree of ipsilesional thalamic atrophy relative to normal, and control for the effects of age, sex, and intracranial volume (ICV), normative deviation values (Z-scores) were calculated using the CentileBrain subcortical model (Ge et al., 2024; centilebrain.org), based on over 37,000 healthy individuals from across the lifespan. Absolute estimates of thalamus volume were not available from the control sample for comparison. We opted not to compute deviation values based on the contralesional side instead, since it has occasionally been reported to show both thalamus volume abnormalities (De Reuck et al., 1995; Hervé et al., 2005) and electrophysiological abnormalities after stroke (e.g. Ahmed, 1988; Johnston et al., 2023; Machado et al., 2004; Tecchio et al., 2007; Wu et al., 2016) suggesting the contralesional side may not be suitable as a comparator overall.

### Experimental design and statistical analysis

Partial Least Squares Correlation (PLSC) was the primary tool for statistical analysis in this work (Krishnan et al., 2011; McIntosh et al., 1996). PLSC is a statistical technique capable of extracting multivariate patterns of correspondence, called latent variables (LVs) between a matrix of high-dimensional brain data (M1) and a matrix containing a set of additional variables of interest (M2), typically design or behavioural variables. Computed by singular value decomposition (SVD) of the covariance matrix of M1 and M2, the resulting LVs comprise a set of “saliences” (eigenvectors), each with an associated “singular value” (square root of the eigenvalue). Saliences quantify the involvement of each element of M1 and M2 (e.g. electrode, voxel, behavioural variable, etc.) in the LV (analogous to PCA loadings), while singular values are proportional to the variance explained by that LV. The PLSC implementation used here has the advantage of assessing the statistical significance and reliability of the LV as a whole (via permutation and bootstrap resampling, respectively) without the need for multiple comparisons correction over matrices with many elements (McIntosh & Lobaugh, 2004). Since PLSC requires an equal number of observations in each row of the input matrices, a PCA-based imputation method (MissMDA; Josse et al., 2011) was used to replace missing values where necessary. The percentage of required imputations for each analysis is reported with the results below, where applicable.

In addition to PLSC, independent samples t-tests were used to assess univariate differences between groups or hemispheres where applicable, and single sample t-tests were used to test for differences against *a priori* values where necessary. Causal Mediation Analysis (CMA, Mediation v5.4.0; Tingley et al., 2014) was used to confirm whether reduced model-estimated intrathalamic inhibitory feedback (Z) mediates the relationship between thalamus degeneration and spectral slowing. A two-tailed significance threshold of α = 0.05 was used across all analyses.

In all reported analyses where thalamus volume was included as an independent variable, normative deviation values (corrected for age, sex and ICV) were used. Overall results did not differ when using uncorrected thalamus volume instead, except where noted (see mediation analysis results below).

## Results

### Lesion and impairment characteristics

Thirteen patients had right hemisphere lesions and twelve had left hemisphere lesions, and the overall mean lesion size was 33.44 cm^3^ (SD = 41.42 cm^3^). Comparing lesion masks to the vascular territory atlas in MNI space (Liu et al., 2023) revealed that lesions were predominantly located in the middle cerebral artery (MCA) territory in this sample (Figure 1), although there was some minor overlap with anterior cerebral artery (ACA) and posterior cerebral artery (PCA) territories.

**Figure 1:**
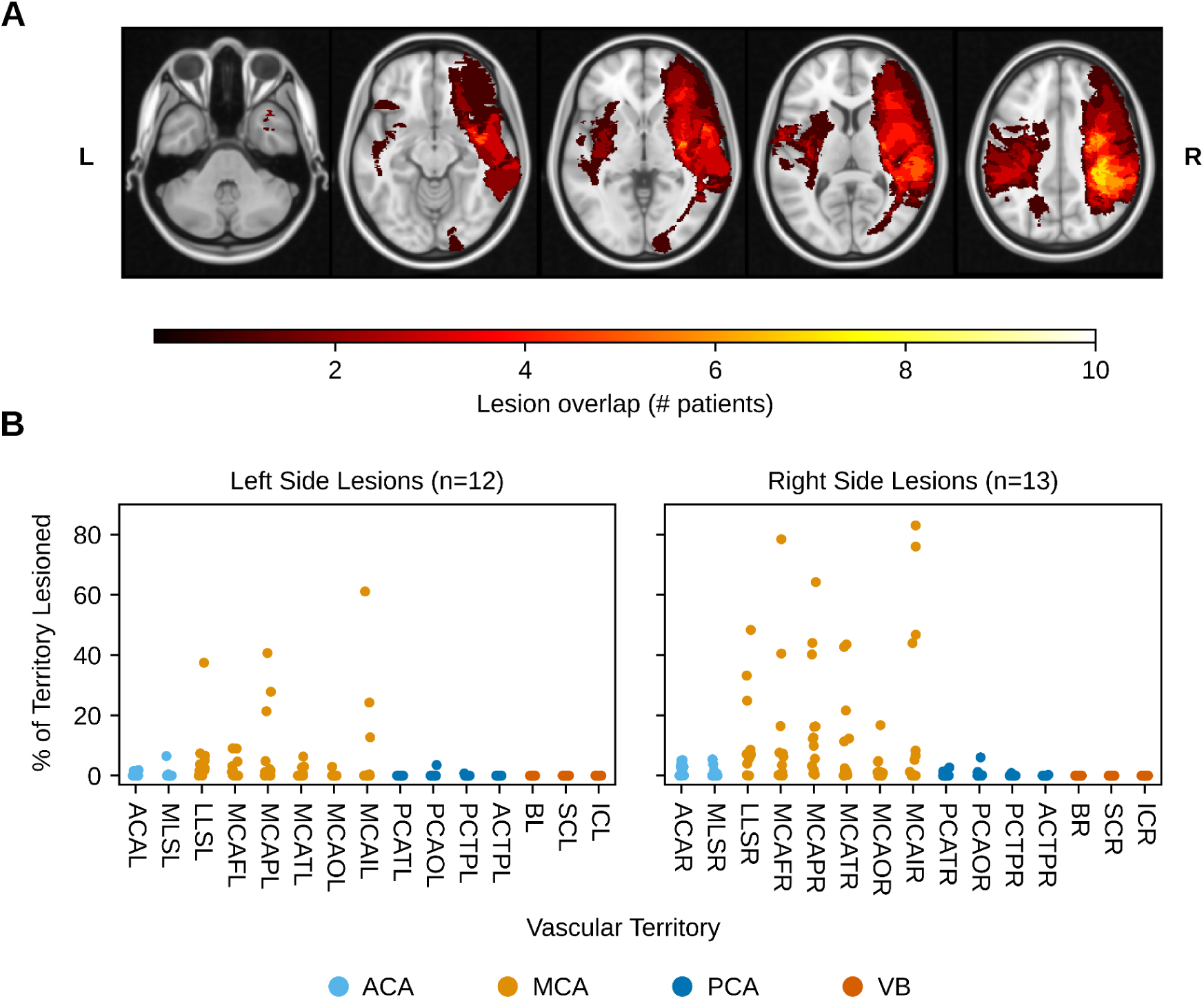
Lesion characteristics. Lesions were primarily concentrated in the vascular territory supplied by the middle cerebral artery (MCA). (A) Heatmap depicting the distribution of lesioned voxels in MNI space. (B) Percentage of each minor vascular territory lesioned, as described in Liu et al. (2023). Colours denote the major vascular territories: ACA = anterior cerebral artery, MCA = middle cerebral artery, PCA = posterior cerebral artery, VB = vertebro-basilar. Minor vascular territory abbreviations are listed in Supplementary Information.

Patients had an average UEFM score of 43.8 out of 66 (SD = 12.74), indicating moderately severe motor impairment overall (higher is better; Duncan et al., 1994), and an average NIHSS score of 2.76 (SD = 2.05) out of a possible 42 (lower is better).

### Stroke patients exhibit broad spectral slowing and model-estimated thalamus disinhibition compared to controls

As a group, stroke patients showed broad elevation of low frequency power, reduction in beta power, and lower alpha frequency compared to controls. Figure 2 summarizes the differences over the entire ipsilesional and contralesional hemispheres (median over all electrodes within a hemisphere, 85 electrodes per hemisphere, midline excluded), while Figure 3 depicts electrode-level statistics.

**Figure 2:**
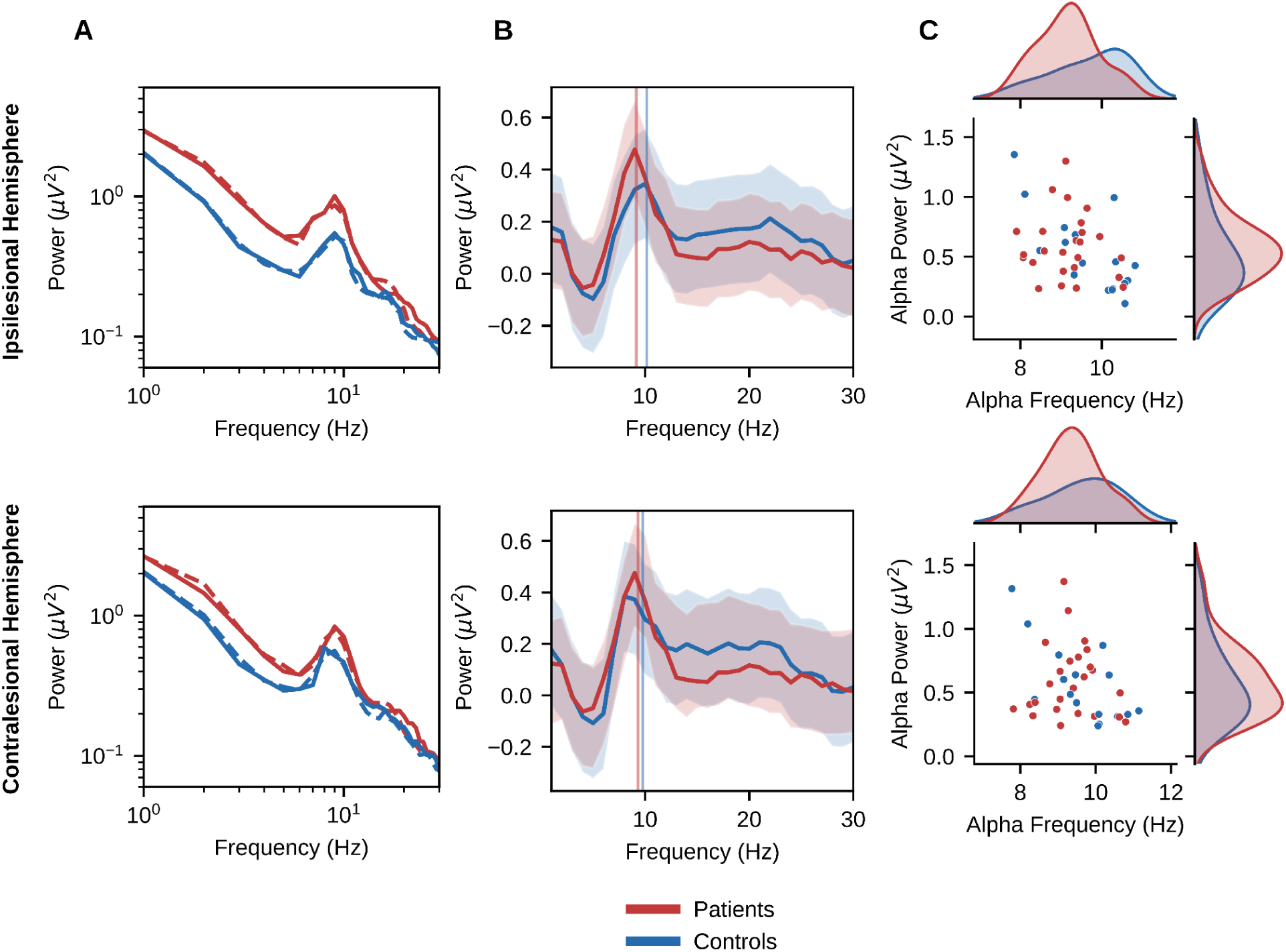
Spectral slowing in stroke patients. Taking the median over all electrodes (within each hemisphere) and participants, patients show the expected features of spectral slowing (electrode-level statistics are in Figure 3). (A) As a group, patients exhibited elevated low frequency power compared to controls over both hemispheres. The simulated power spectra (dashed lines) from the corticothalamic model accurately reproduced this feature. (B) Power spectra with aperiodic component subtracted, suggesting alpha slowing and beta power reduction in patients. Vertical lines show the median estimated alpha frequency. (C) Spectral parameterization (*specparam)* explicitly quantifies alpha frequency, suggesting alpha slowing in patients.

**Figure 3:**
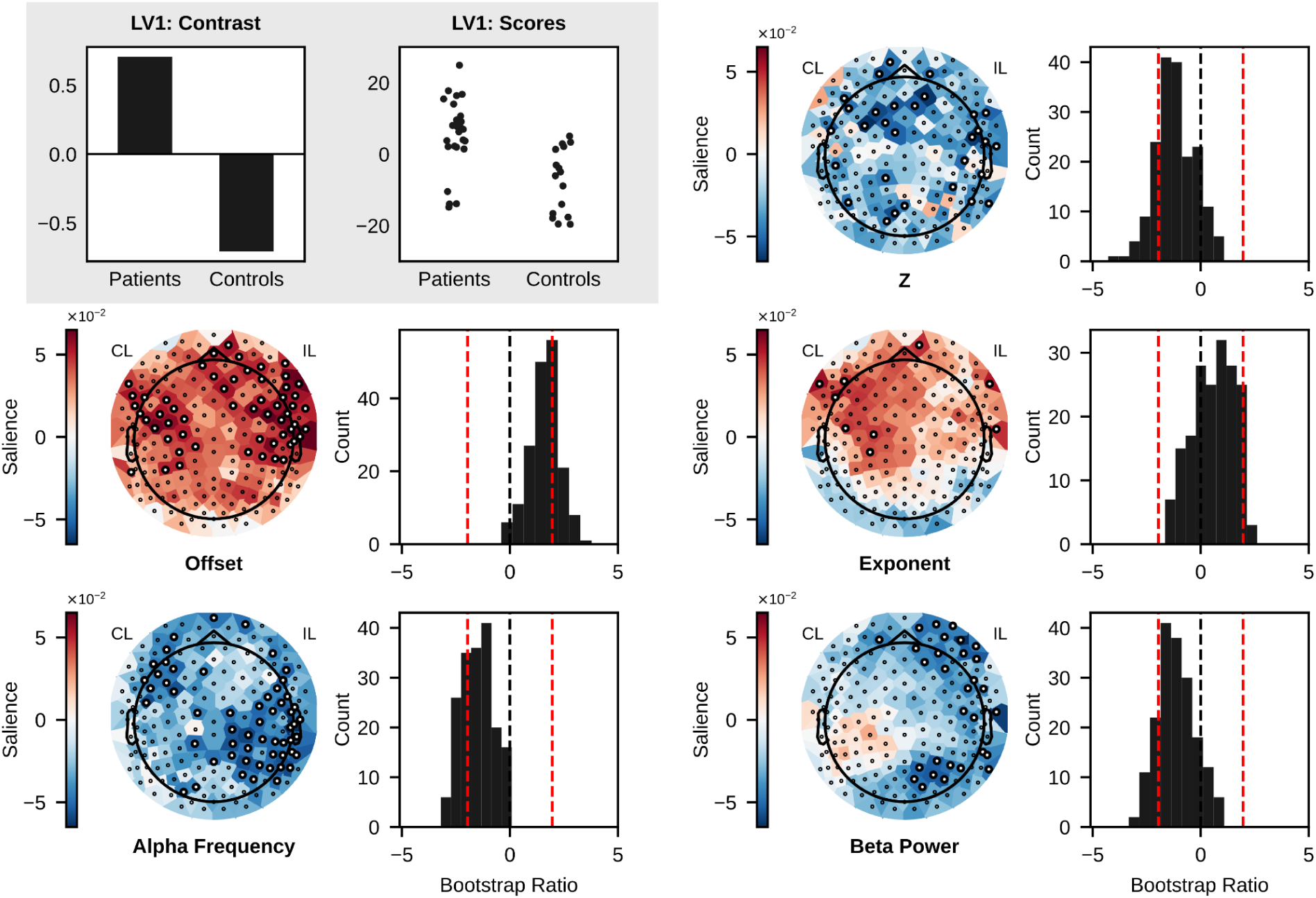
Mean-centered PLSC contrasting patients and controls. Patients showed the predicted abnormalities in spectral parameters and fitted corticothalamic model parameters. This figure depicts the first and only significant latent variable (LV1): s=9.09, p = 0.026. Grey box: PLSC contrast (Left) and scores (Right). The contrast indicates that patients are associated positively with the topographies depicted in the other plots, and controls are associated with the inverse pattern. Scores indicate the degree to which individual patients and controls express the depicted pattern. Paired plots: Each pair of plots depicts saliences over electrodes (Left) and bootstrap ratios (Right) for the given parameter. Saliences quantify the degree to which a given parameter at a given electrode differentiates patients and controls, following the contrast coding in the top left. The valence (positive or negative) of the salience indicates whether that correspondence is positive or negative. Bootstrap ratios quantify the reliability of this correspondence at a given electrode via bootstrap resampling, where an absolute value greater than 1.96 has a probability of 0.05 given a unit normal two-tailed distribution (these electrodes are highlighted). Bootstrap ratios of 0 and ±1.96 are marked on the histograms by dashed black and red lines, respectively.

These stroke-related power spectrum abnormalities were satisfactorily parameterized by *specparam*, as fits were good for patients (mean R^2^ = 0.94, SD = 0.05), and controls (mean R^2^ = 0.93, SD = 0.06), with no significant difference between the groups (t(40) = 0.52, p = 0.610). Similarly, *BrainTrak* model fits were good overall for patients (mean likelihood = 0.88, SD = 0.02) and controls (mean likelihood = 0.87, SD = 0.02), with no significant difference between groups (t(40) = 1.59, p = 0.121). Average empirical and simulated power spectra are shown in Figure 2A, demonstrating the ability of the corticothalamic model to reproduce the group-level abnormalities in the stroke patient power spectra.

To test the predicted group differences in the resulting spectral parameters (exponent, alpha frequency, and beta power) and model-estimated intrathalamic inhibition (Z), we used mean-centered PLSC (McIntosh & Lobaugh, 2004) at the electrode level, which also reveals the spatial distribution of these differences over the scalp (Figure 3). While our previous work with MEG had treated the aperiodic offset as redundant with the aperiodic exponent, here we opted to also include aperiodic offset in the PLSC brain matrix to clarify any differences in the magnitude and topography of these two closely related parameters. 6.8% of the PLSC brain matrix was imputed due to missing values.

PLSC extracted one significant LV that differentiated patients from controls (Figure 3, LV1: s = 9.091, p = 0.026). All predictions with respect to spectral slowing were supported. Namely, aperiodic offset and exponent were both elevated broadly across the scalp, although offset showed a somewhat more widespread and reliable difference. Conversely, alpha frequency and beta power were widely decreased in patients, as observed in previous work (Johnston et al., 2023). Finally, patients exhibited a widespread deficit in the corticothalamic model parameter Z, indicating the predicted thalamic disinhibition compared to controls. This pattern of PLSC results was largely unchanged when the brain matrix was first residualized by age (Supplementary Figure S2).

### Patients show thalamus atrophy

Consistent with our previous results, patients showed substantial thalamic atrophy as a group, particularly in IL thalamus (Figure 4). Raw IL thalamus volume (mean = 3250 mm^3^, SD = 796 mm^3^) was significantly smaller than CL thalamus volume (mean = 4623 mm^3^, SD = 857 mm^3^; t(24) = -7.59, p << 0.001). Normative deviation values were also significantly lower for IL thalamus (mean = -6.75, SD = 1.98) than for CL thalamus (mean = -4.30, SD = 1.28; t(24) = -7.75, p << 0.001). Estimates of thalamus volume did not differ significantly between raters for the IL thalamus volume (t(23) = -0.81, p = 0.424), CL thalamus (t(23) = 0.16, p = 0.876) or ratio of IL to CL thalamus volume (t(23) = -0.89, p = 0.381).

**Figure 4:**
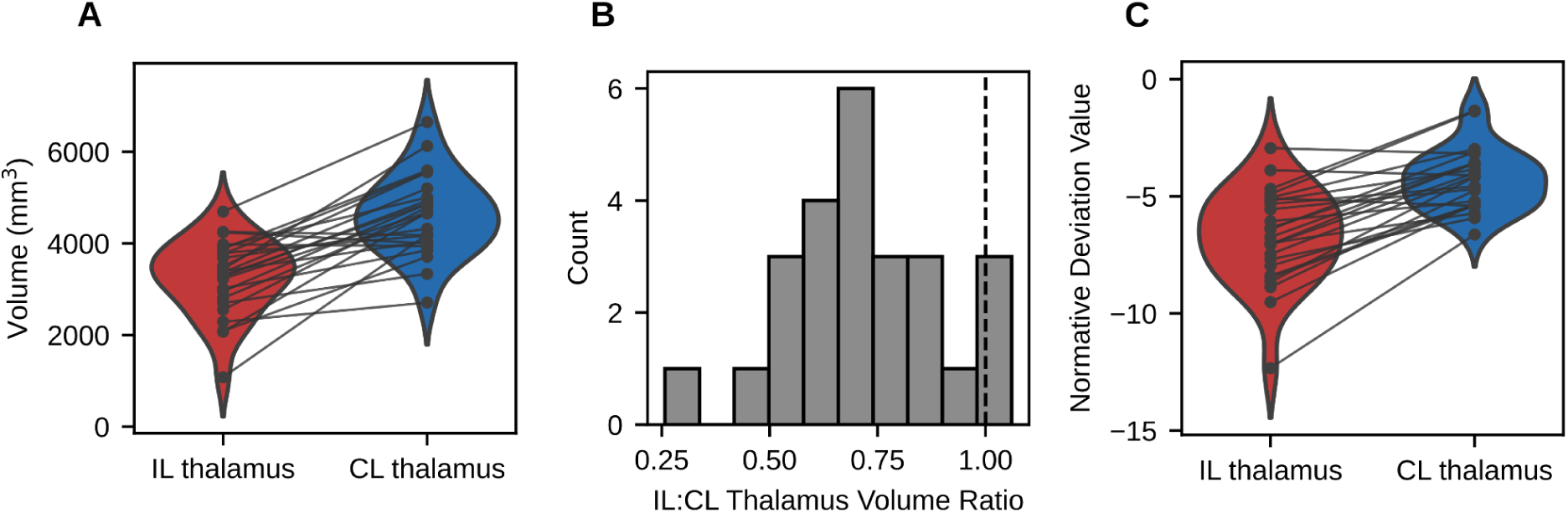
Thalamus volume comparison. Stroke patients showed substantial thalamus atrophy. (A) Absolute volume of ipsilesional (IL) thalamus versus contralesional (CL) thalamus. (B) Distribution of IL:CL thalamus volume ratios. The dashed black line marks a ratio of 1 (i.e. no difference). (C) Normative deviation values of ipsilesional (IL) thalamus versus contralesional (CL) thalamus.

### Thalamus degeneration is related to spectral slowing and model-estimated thalamic disinhibition

Given the observed atrophy in IL thalamus, we used PLSC to verify that this atrophy was related to spectral slowing and model-estimated thalamic disinhibition. In addition to ipsilesional thalamus volume (corrected for age, sex, and ICV as noted above), we also included some additional predictor variables in the model: infarct volume (residualized by ICV) to account for overall stroke severity, as well as time post onset (TPO) to account for potential variation related to time elapsed since the stroke. We also included UEFM and NIHSS scores to explore whether behavioural impairments covaried with the extracted patterns. 7.2% of the PLSC brain matrix was imputed due to missing values.

Consistent with previous findings, PLSC extracted a significant LV relating IL thalamus volume to a broad pattern of spectral slowing, and moderate model-estimated thalamic disinhibition (Figure 5; LV1: s = 11.65, p < 0.001, variance explained = 0.56). Specifically, IL thalamus volume had the highest salience and correlation (R) values and was therefore most strongly implicated in the LV (salience = 0.80; R = 0.67, 95% CI [0.62, 0.86]), followed by the infarct volume (salience = -0.54; R = -0.45, 95% CI [-0.21, -0.71]). None of the other predictor variables had large saliences, or correlations that were significantly different from zero: TPO (salience = 0.041; R = 0.034, 95% CI [-0.34, 0.30]), UEFM score (salience = 0.21; R = 0.17, 95% CI [-0.12, 0.56]), and NIHSS score (salience = 0.18; R = 0.15, 95% CI [-0.22, 0.46]).

**Figure 5:**
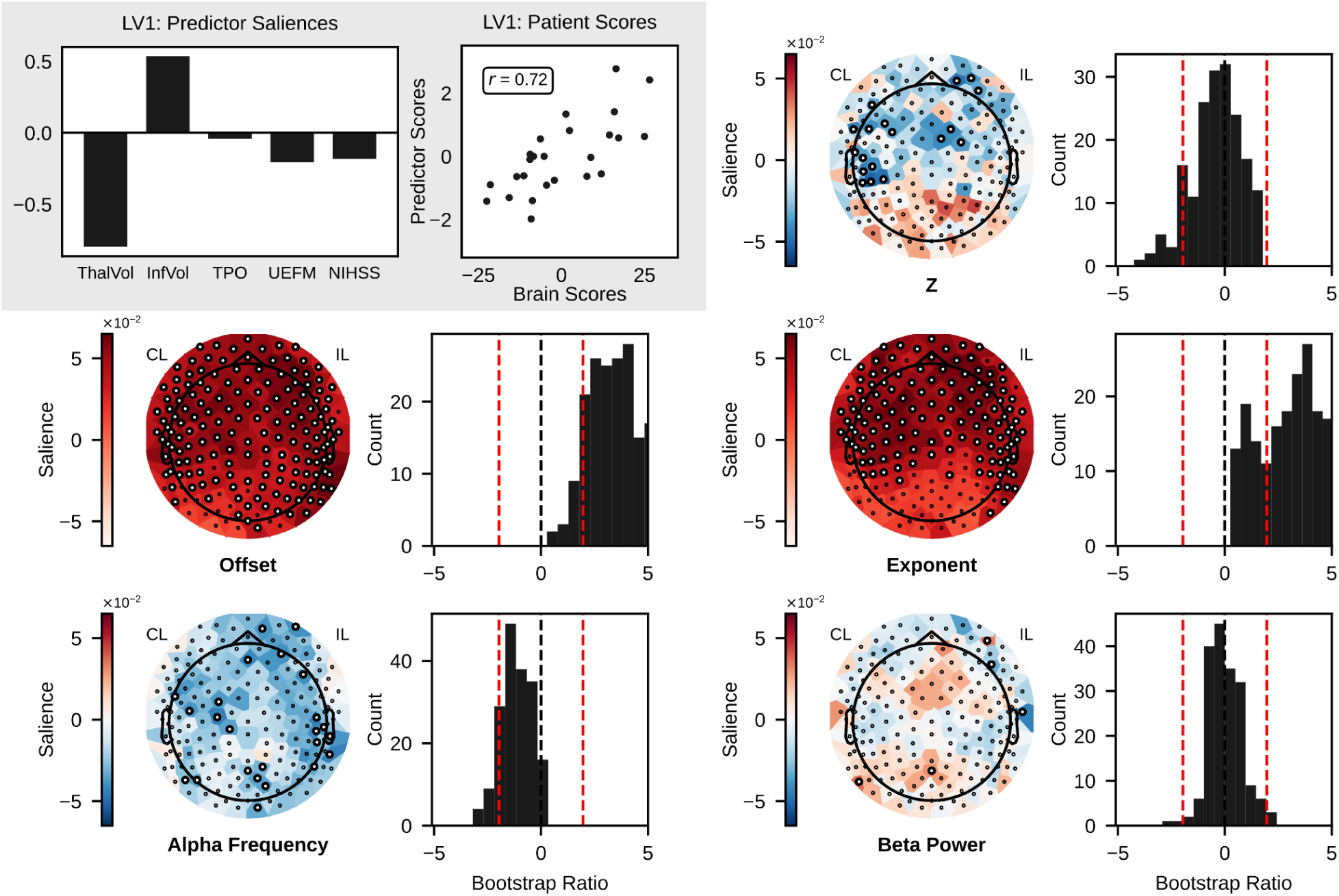
PLSC relating abnormal spectral and corticothalamic model parameters to thalamus atrophy and other predictors. Patients demonstrated the expected pattern, where thalamic atrophy was associated with decreased intrathalamic inhibitory feedback (Z) and spectral parameters mostly indicative of spectral slowing (higher offset, higher exponent, lower alpha frequency, but no beta power deficit). This figure depicts the first and only significant latent variable (LV1): s = 11.65, p < 0.001, variance explained = 0.56. Grey box: Saliences for the five included predictors (Left) and predictor scores versus brain scores for each patient (Right). Larger saliences on the left indicate a stronger correspondence between the predictor and the brain salience topographies depicted in the other plots. Whiskers denote the 95% confidence interval as determined by bootstrap resampling. ThalVol: ipsilesional thalamus volume, InfVol: infarct volume, TPO: time post onset, UEFM: Upper Extremity Fugl-Meyer (total score), NIHSS: National Institutes of Health Stroke Scale (total score). Predictor scores indicate the degree to which an individual patient expresses the pattern of predictor saliences on the left, while brain scores indicate the degree to which an individual patient expresses the pattern of brain saliences depicted in the topography plots. Paired plots: Each pair of plots depicts saliences over electrodes (Left) and bootstrap ratios (Right) for the given parameter. Saliences quantify the correspondence between a given parameter at a given electrode and the pattern of predictor variables extracted by the LV shown in the top left, and the valence (positive or negative) indicates whether this correspondence is positive or negative. Bootstrap ratios quantify the reliability of this correspondence at a given electrode via bootstrap resampling, where an absolute value greater than 1.96 has a probability of 0.05 given a unit normal two-tailed distribution (these electrodes are highlighted). Bootstrap ratios of 0 and ±1.96 are marked on the histograms by dashed black and red lines, respectively.

Regarding the brain variables, aperiodic exponent (and offset) showed broadly positive saliences, particularly over anterior scalp, while alpha frequency showed negative saliences over nearly the entire scalp. Of the spectral parameters previously associated with spectral slowing, only beta power did not show a clear pattern of negative saliences in relation to the predictor variables. Importantly, Z (intrathalamic inhibitory feedback) also showed negative saliences, predominantly over anterior scalp, replicating the previously observed association between thalamus degeneration and model-estimated thalamus disinhibition.

In this analysis thalamus volume, the predictor of primary interest, was statistically corrected for age, but we also considered the role of age directly by including it as a predictor (Supplementary Fig S2). In that case, thalamus volume was still the strongest predictor of EEG abnormalities, but age also loaded heavily on the LV. The resulting brain pattern heavily emphasized offset and exponent across the scalp, and de-emphasized alpha slowing and Z compared to when age was not included.

### Estimated thalamic disinhibition does not significantly mediate the relationship between thalamic atrophy and spectral slowing

We used Causal Mediation Analysis (Mediation v5.4.0; Tingley et al., 2014) to test whether reduced model-estimated intrathalamic inhibitory feedback (reduced Z) links thalamic degeneration and EEG spectral slowing. Specifically, we used each individual patient’s ipsilesional thalamic volume deviation as the independent variable, Z as the mediator, and a combined measure of spectral slowing as the dependent variable. This spectral slowing measure was computed as a weighted sum of spectral parameters, with the weightings taken from our previous work in MEG (Johnston et al., 2023, 2024; exponent = 0.63, alpha frequency = -0.67, beta power = -0.38). Spectral parameters were Z-scored, then multiplied by their weights and summed for each patient.

The direct effect of ipsilesional thalamus volume deviation on spectral slowing was statistically significant when averaging over the entire ipsilesional hemisphere (estimate = -0.41, p = 0.02), as well as frontal (estimate = -0.36, p = 0.03), prefrontal (estimate = -0.29, p = 0.03), central (estimate = -0.38, p = 0.03), and temporal (estimate = -0.40, p = 0.01) electrode groups (see Supplementary Table S1 for electrode groupings). However, contrary to prior work, the mediation effect was not significant when averaging over the ipsilesional hemisphere (proportion of total effect via mediation = 0.05, p = 0.72), nor over individual electrode groups (proportion of total effect via mediation = -0.03 to -.25, p = 0.142 to 0.88). We note, however, that partial mediation effects were found at prefrontal, frontal, and central electrode groups when using uncorrected ipsilesional thalamus volume as the independent variable (Supplementary Table S2).

### Thalamus atrophy is not significantly correlated with available behavioural outcome measures

To assess whether thalamus degeneration was associated with the available behavioural measures, we computed Spearman correlations between ipsilesional thalamus volume deviation values and UEFM and NIHSS scores (Figure 6). To account for the possible shared effect of overall stroke severity on both thalamus volume and the behavioural measures, correlations were computed on the residuals after regressing both independent and dependent variables on infarct volume. Residualized ipsilesional thalamus volume deviation was not significantly correlated with the UEFM residuals (*r_s_*(23) = 0.33, p = 0.1), nor with the NIHSS residuals (*r_s_*(23) = 0.30, p = 0.14).

**Figure 6:**
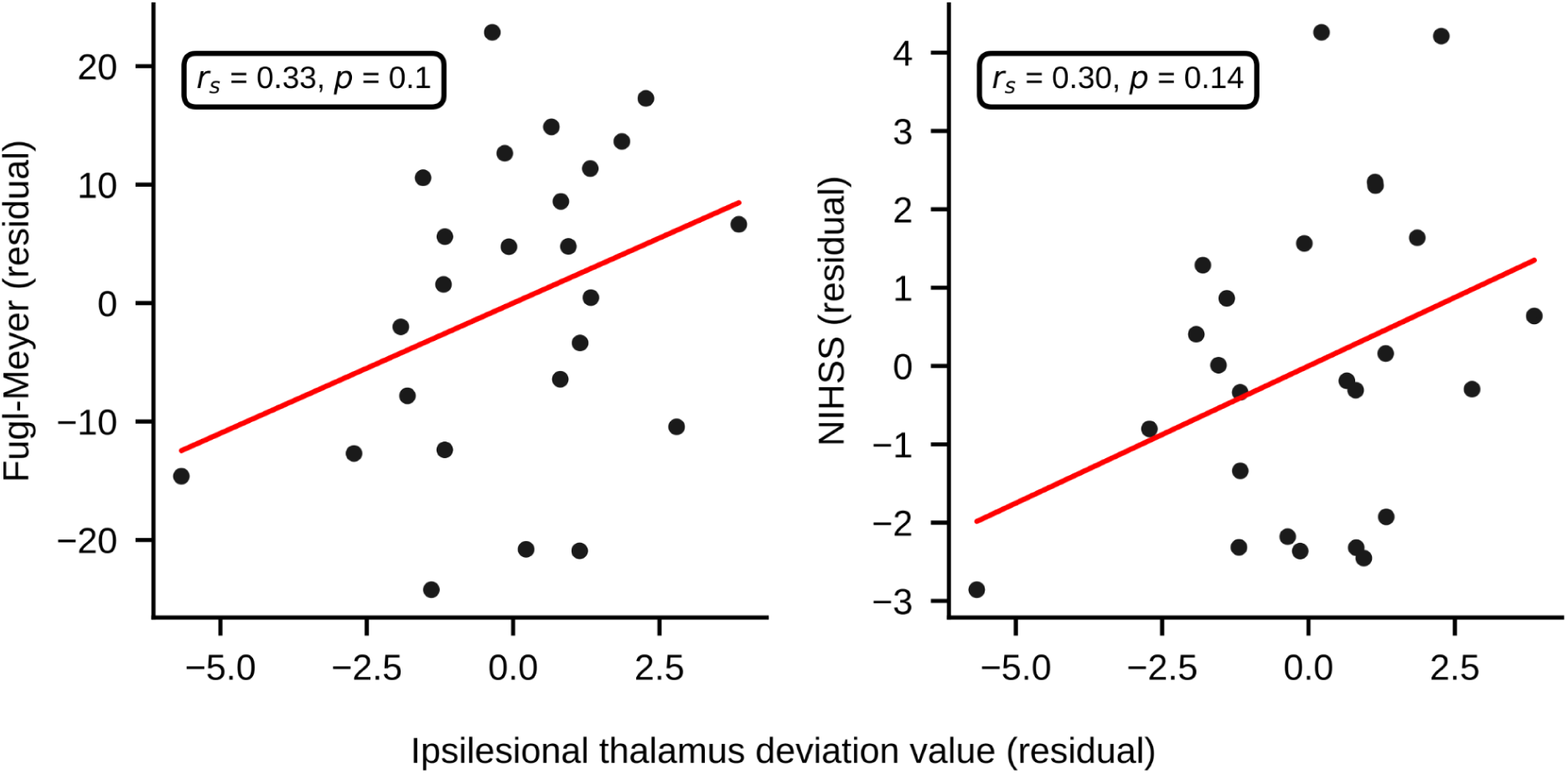
Behavioural measures and ipsilesional thalamus volume normative deviation values after correcting for infarct volume. The available behavioural measures, Upper Extremity Fugl-Meyer and NIHSS, were not significantly related to ipsilesional thalamus volume. Reported statistics are Spearman correlations computed on the residuals after regressing both independent and dependent variables on infarct volume.

## Discussion

In this work, we further clarified the nature of post-stroke neural slowing, and found corroborating evidence linking this slowing to secondary degeneration of the thalamus.

First, stroke patients showed the previously identified abnormalities after spectral parameterization, i.e. increased aperiodic exponent, decreased alpha frequency, and decreased beta power (as well as a higher aperiodic offset). This further establishes that the classic large scale neural slowing phenomenon, detected in dozens of datasets over the past several decades, is largely attributable to changes in the aperiodic background activity, rather than low frequency oscillations, although genuine oscillatory abnormalities in the alpha and beta bands were also present (Johnston et al., 2023).

Patients also demonstrated the expected correlational evidence of secondary thalamus dysfunction on the ipsilesional side, including reduced intrathalamic inhibitory feedback (Z) as estimated by *BrainTrak*, and reduced thalamic volume (Johnston et al., 2024). Crucially, PLSC revealed the expected relationships between abnormal neural dynamics and thalamus volume deviation (i.e. thalamus volume corrected for age, sex, and ICV), where reduced ipsilesional thalamus volume was associated with higher aperiodic exponent (and offset), lower alpha frequency, and lower Z. Furthermore, thalamus volume was a substantially stronger predictor of these features than overall lesion size or time since stroke onset, despite the absence of direct thalamus injury in these patients, further supporting the role of secondary thalamus degeneration in large-scale neural dysfunction post-stroke. We note, however, that beta power did not show a clear relationship with thalamus volume, despite being significantly reduced in patients compared to controls, suggesting that its causes may differ from the aperiodic abnormalities and alpha slowing.

While corroborating these prior findings, this study also offers several new insights. First, this work demonstrates that secondary thalamic degeneration and related electrophysiological abnormalities can occur in patients presenting with motor impairment, and can be caused by a stroke, in either the right or left hemisphere, that did not directly damage the thalamus. From a research translation perspective, it is also notable that the primary effects of interest, previously observed with MEG multimodal MRI, were replicable here with sensor-level EEG data and structural MRI alone, given the relative accessibility of these techniques in both clinical and research settings. Furthermore, the multivariate, sensor space analyses used here provide new insight into the topography of abnormal neural dynamics, indicating that the spectral features associated with thalamus degeneration, while correlated, have different spatial extents. Specifically, aperiodic exponent and offset tended to be more abnormal over anterior scalp, while alpha slowing was spread more evenly across the scalp. The anterior topography of the aperiodic abnormalities is notable given previous work showing a relationship between thalamic neuronal loss and hypometabolism in frontal cortex (Yamauchi et al., 2022). Moreover, these group-level topographies were quite symmetrical across the midline, underscoring the finding that ipsilesional thalamic atrophy is associated with bilateral dysfunction, although volume conduction may have obscured some differences in magnitude between the hemispheres.

While our predictions were supported overall, there are some important caveats and limitations to note. First, evidence that thalamic disinhibition (lower Z) mediates the relationship between thalamus volume deviation and large-scale neural slowing did not pass a two-tailed significance level of 0.05 when variables were averaged over the whole ipsilesional hemisphere, nor over smaller electrode clusters (however, marginal mediation effects were detected when using raw thalamus volume scores as the independent variable, Supplementary Table S2). Similarly, PLSC identified the expected relationship between thalamus volume deviation and Z, but it was substantially weaker than the relationship with neural slowing (Figure 5). On the other hand, the direct effect linking thalamus atrophy to neural slowing was significant across much of the scalp. As such, these findings support the hypothesis that secondary thalamus degeneration is related to neural slowing, but evidence to support thalamic disinhibition as the causal mechanism linking thalamic degeneration to neural slowing was limited compared to prior work (Johnston et al., 2024). However, we note some limitations of the data that may have contributed to this ambiguity. For instance, heterogeneity in lesion location and size, while important for establishing generalizability, may have obscured more subtle mediation effects, as not all individuals would express the effects consistently at the same electrodes - an issue potentially compounded by slight variations in electrode placement between individuals. Additionally, estimates of thalamus degeneration could have been less reliable here since only one coarse measure (thalamus volume deviation) was used, rather than the combination of three used previously (thalamus volume, mean diffusivity, and blood flow; Johnston et al., 2024). Similarly, EEG suffers from more contamination above 15 Hz than beamformed MEG, which may have reduced the reliability of parameter estimation via *BrainTrak* and *specparam*, potentially obscuring the interrelationships among spectral parameters, Z, and thalamus atrophy. Despite these caveats, we conclude that large-scale neural slowing remains a promising biomarker for indexing neurophysiology underlying post-stroke brain dysfunction - one that is objective, readily measurable with EEG or MEG, and biologically grounded (Wang, 2026). However experimental work is still necessary to directly validate the disease mechanisms underlying secondary thalamic injury.

Furthermore, we did not find a significant relationship between thalamus volume deviation and the available measures of behavioural impairment (UEFM and NIHSS) indicating that thalamus degeneration may not be a primary determinant of the motor and neurological impairments captured by these measures. However, severely neurologically impaired participants were underrepresented in the sample, and overall variance was low (mean NIHSS score: 2.76 out of 42, SD: 2.05), potentially limiting our ability to detect a relationship between neurological impairment and thalamus degeneration. Other cognitive and language assessments were not available for these patients, so the previously observed effect of thalamus degeneration on these domains could not be assessed (Geng et al., 2023; Johnston et al., 2024).

Additionally, the effect of age needs to be considered, as the sample of stroke patients was younger than the controls on average due to necessary exclusions in both groups. However, this imbalance only directly affects the group comparisons, which did not differ from previous work, regardless of whether age was first regressed from the independent and dependent variables (Figure 3; Supplementary Figure S2). While age has been previously associated with lower aperiodic exponents (Voytek et al., 2015), an independent analysis of a partially overlapping dataset with the one used here found a significant moderating effect of age, with older stroke patients instead showing higher aperiodic exponents (Albertson et al., 2025). In our investigation, while thalamus atrophy remained the strongest predictor overall, our supplementary PLSC analysis (Supplementary Figure S3) identified a correspondence between younger age, thalamus atrophy, higher aperiodic exponent and higher aperiodic offset. Notably, neither work found a relationship between alpha slowing and age, suggesting that age may moderate the aperiodic aspect of neural slowing but not necessarily the alpha oscillatory component. Overall, these ambiguities highlight the need for larger, ideally longitudinal datasets to disentangle the effects and interactions of critical variables like age, thalamus degeneration and infarct volume on large-scale neural dynamics.

Finally, it should be noted that the majority of patients in the current sample had infarcts within the MCA territory (Figure 1). This is expected given the predominance of MCA strokes compared to those involving other major vascular territories (Hossmann & Heiss, 2010), but we cannot assume that these findings apply to patients with other types of stroke. Future work may need to deliberately recruit patients with non-MCA strokes to determine whether, and to what extent, they are affected by secondary thalamic degeneration.

In conclusion, this study has corroborated a more refined view of the classic finding in stroke electrophysiology, recasting neural slowing as a conjunction of abnormalities in the aperiodic exponent, aperiodic offset, alpha slowing, and beta power. It has also further established a link between these abnormalities (with the exception of beta power reduction) to secondary injury of the thalamus. Together, these findings further clarify the nature and extent of abnormal neural dynamics in patients with stroke in the MCA territory, and establish secondary thalamic degeneration as an important effect of stroke with significant implications for post-stroke brain dysfunction. In doing so, this work helps bridge the gap between the large existing stroke electrophysiology literature on neural slowing, and the underlying disease processes - a crucial step towards useful biomarkers that can reduce the burden of stroke.

## Data Availability

Raw data is available upon reasonable request to Dr. Cramer.

## Author contributions

Johnston, P.R.: Conceptualization, Methodology, Software, Formal Analysis, Data Curation, Writing - Original Draft, Visualization

Schwarz, A.: Data Curation, Writing - Review & Editing

Dodd, J.: Formal Analysis, Data Curation

Griffiths J. D.: Conceptualization, Methodology, Software, Writing – Review & Editing.

Meltzer, J. A.: Conceptualization, Methodology, Writing – Review & Editing

Cramer, S. C.: Conceptualization, Resources, Project Administration, Writing - Review & Editing

McIntosh, A. R.: Conceptualization, Methodology, Writing – Review & Editing, Supervision

No generative artificial intelligence tools were used to prepare this manuscript.

## Acknowledgements

P.R.J.: Natural Sciences and Engineering Research Council of Canada (NSERC) CGS-D

J.D.G.: Krembil Foundation, CAMH Discovery Fund, Labatt Family Network, UToronto EMHSeed

J.A.M.: Canada Research Chair, NSERC Discovery Grant.

S.C.C: National Institutes of Health (K24HD074722)

A.R.M.: NSERC Discovery Grant

This research was enabled by computing resources provided by Calcul Québec (calculquebec.ca) and the Digital Research Alliance of Canada (alliancecan.ca).

