## Supplementary Material for "Large-scale neural slowing measured with electroencephalography indexes secondary thalamic degeneration in stroke"

**Figure 1 caption continued**: Minor territory abbreviations (last letter denotes left/right): ACA = anterior cerebral artery, MLS = medial lenticulostriate, LLS = lateral lenticulostriate, MCAF = frontal pars of middle cerebral artery, MCAP = parietal pars of middle cerebral artery, MCAT = temporal pars of middle cerebral artery, MCAO = occipital pars of middle cerebral artery, MCAI = insular pars of middle cerebral artery, PCAT = temporal pars of posterior cerebral artery, PCAO = occipital pars of posterior cerebral artery, PCTP = posterior choroidal and thalamoperforators, ACTP = anterior choroidal and thalamoperforators, B = basilar, SC = superior cerebellar, IC = inferior cerebellar.

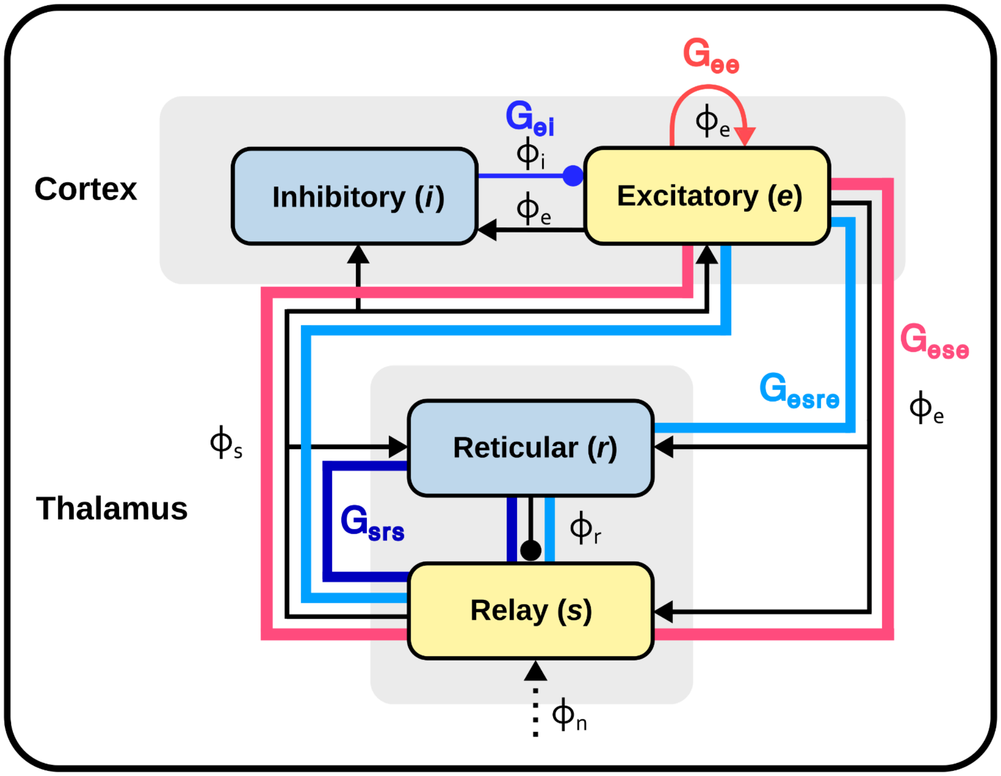


**Figure S1**: The neurophysiological model of the corticothalamic system used to simulate power spectra. This model emulates the interactions among two cortical neuronal populations (one inhibitory [i] and one excitatory [e]) and two thalamic neuronal populations (the excitatory relay nuclei [s] and inhibitory reticular nucleus [r]). Excitatory populations are represented by yellow boxes, and inhibitory populations by blue boxes. This model is described by five gain parameters ($G_{ee}$, $G_{ei}$, $G_{ese}$, $G_{esre}$, $G_{srs}$) representing the ratio of input to output within key loops of the circuit, as well as two parameters shared by all four populations (α: synaptic decay time, β: synaptic rise time) and two additional parameters ($t_0$: thalamocortical delay, $A_{EMG}$:

electromyographic artifact correction). Φ symbols represent the axonal signals propagating from their respective neuronal populations, with the simulated power spectrum being derived from $\Phi_e$. These parameters can be further aggregated into three simplified parameters X, Y, and Z, which summarize the gain of the three major loops: intracortical, corticothalamic, and intrathalamic, respectively. See Robinson et al., (2001), Abeysuriya & Robinson (2016), and Supplementary Information from Johnston et al. ( 2024) for more information.

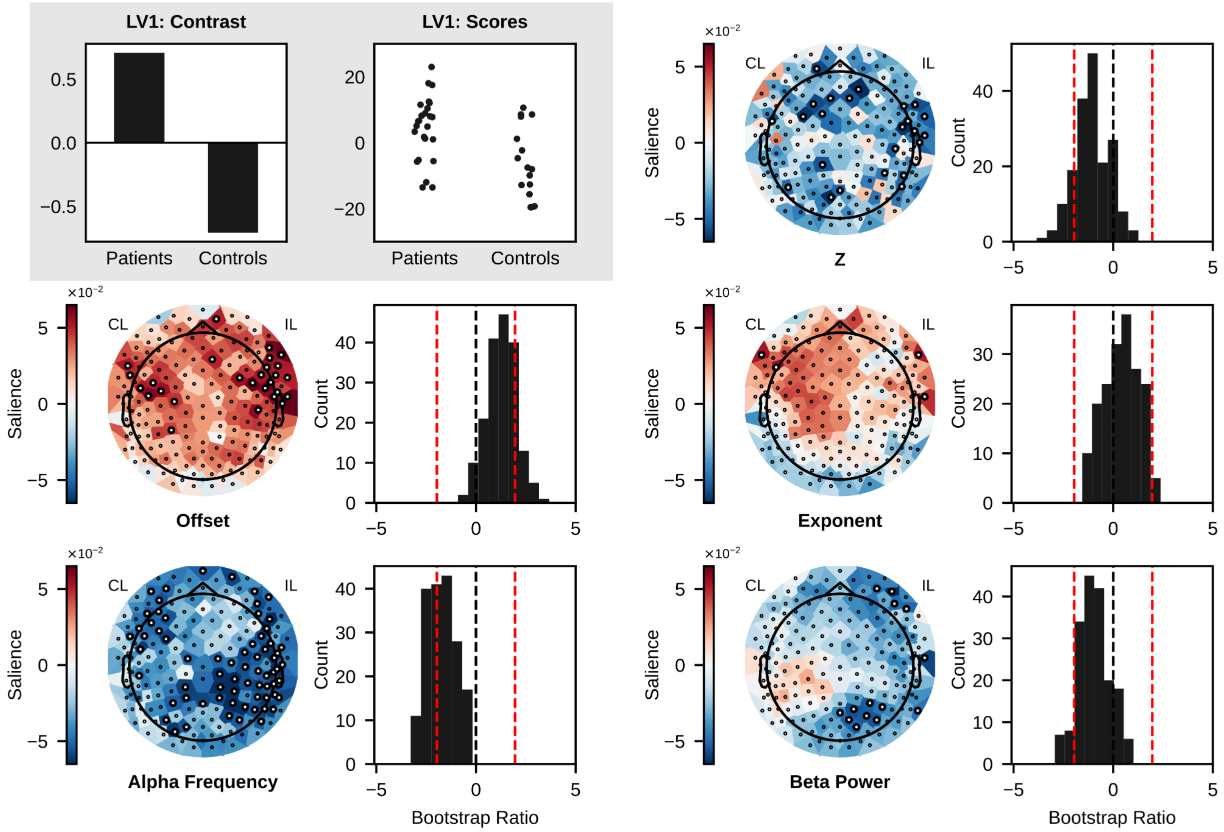


**Figure S2**: Figure 3 PLSC after residualizing brain matrix by age, p = 0.06, s = 8.473.

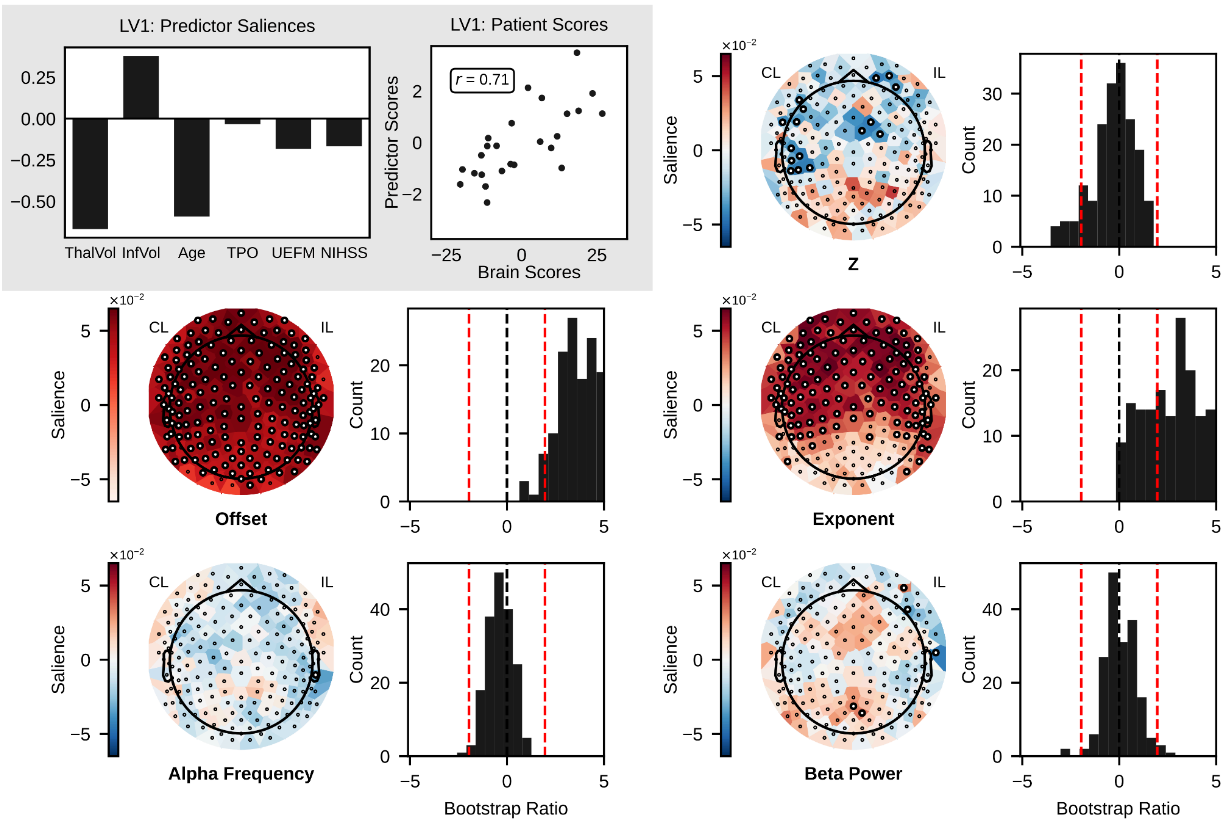


**Figure S3**: Figure 5 with age added as a predictor. LV1: $p = 0.002$, $s = 14.027$, variance explained = 0.6.

| | L | R |
|---|---|---|
| Prefrontal | 'E27', 'E33', 'E34', 'E38', 'E39', 'E47', 'E48', 'E37', 'E46', 'E54' | 'E20', 'E19', 'E12', 'E11', 'E3', 'E2', 'E222', 'E18', 'E10', 'E1' |
| Frontal | 'E16', 'E22', 'E23', 'E24', 'E28', 'E29', 'E30', 'E35', 'E36', 'E40', 'E41', 'E42', 'E49', 'E50' | 'E7', 'E14', 'E6', 'E207', 'E13', 'E5', 'E215', 'E4', 'E224', 'E223', 'E214', 'E206', 'E213', 'E205' |
| Central | 'E9', 'E17', 'E43', 'E44', 'E45', 'E51', 'E52', 'E53', 'E57', 'E58', 'E59', 'E60', 'E64', 'E65', 'E66', 'E71', 'E72' | 'E186', 'E198', 'E197', 'E185', 'E132', 'E196', 'E184', 'E144', 'E204', 'E195', 'E183', 'E155', 'E194', 'E182', 'E164', 'E181', 'E173' |
| Temporal | 'E55', 'E56', 'E62', 'E63', 'E69', 'E70', 'E74', 'E75', 'E84', 'E83', 'E95', 'E61', 'E68', 'E94' | 'E221', 'E212', 'E211', 'E203', 'E202', 'E193', 'E192', 'E180', 'E179', 'E178', 'E191', 'E210', 'E220', 'E190' |
| Parietal | 'E76', 'E77', 'E78', 'E79', 'E80','E85', 'E96', 'E86', 'E87', 'E88', 'E89', 'E97', 'E98', 'E99', 'E100', 'E110' | 'E172', 'E163', 'E154', 'E143', 'E131', 'E162', 'E153', 'E142', 'E130', 'E161', 'E152', 'E141', 'E129', 'E128', 'E170', 'E171' |
| Occipital | 'E107', 'E108', 'E109', 'E116', 'E117', 'E118', 'E125', 'E106', 'E115', 'E124', 'E105', 'E114', 'E123', 'E136' | 'E160', 'E151', 'E140', 'E150', 'E139', 'E127', 'E138', 'E149', 'E159', 'E169', 'E177', 'E168', 'E158', 'E148' |

**Table S1**: Electrode region groupings.

| Region | Total Effect | ACME | ADE | Prop. Mediated | $\rho_{ACME=0}$ |
|---|---|---|---|---|---|
| **All** | -0.27<br>[-1.00, 0.12]<br>p = 0.19 | -0.03<br>[-0.29, 0.05]<br>p = 0.45 | -0.25<br>[-0.95, 0.15]<br>p = 0.31 | 0.11<br>[-1.66, 1.89]<br>p = 0.53 | -0.23 |
| **Prefrontal** | -0.47<br>[-0.96, -0.20]<br>p <<0.001 *** | -0.10<br>[-0.32, 0.00]<br>p = 0.058 . | -0.37<br>[-0.79, -0.13]<br>p = 0.004 ** | 0.22<br>[-0.01, 0.58]<br>p = 0.058 . | -0.44 |
| **Frontal** | -0.44<br>[-0.91, -0.14]<br>p = 0.008 ** | -0.11<br>[-0.38, 0.01]<br>p = 0.070 . | -0.33<br>[-0.73, -0.04]<br>p = 0.032 * | 0.26<br>[-0.07, 0.78]<br>p = 0.074 . | -0.49 |
| **Central** | -0.57<br>[-1.12, -0.20]<br>p <<0.001 *** | -0.16<br>[-0.62, 0.02]<br>p = 0.096 . | -0.41<br>[-0.84, -0.10]<br>p <<0.001 *** | 0.29<br>[-0.07, 0.78]<br>p = 0.096 . | -0.62 |
| **Temporal** | -0.36<br>[-0.82, -0.10]<br>p = 0.012 * | -0.05<br>[-0.34, 0.06]<br>p = 0.37 | -0.31<br>[-0.73, -0.06]<br>p = 0.014 * | 0.14<br>[-0.19, 0.74]<br>p = 0.362 | -0.37 |
| **Parietal** | -0.45<br>[-0.97, -0.05]<br>p = 0.022 * | 0.02<br>[-0.27, 0.16]<br>p = 0.886 | -0.46<br>[-0.91, -0.06]<br>p = 0.026 * | -0.04<br>[-0.85, 0.38]<br>p = 0.872 | -0.39 |
| **Occipital** | -0.27<br>[-1.03, 0.14]<br>p = 0.23 | 0.03<br>[-0.18, 0.16]<br>p = 0.77 | -0.30<br>[-1.00, 0.11]<br>p = 0.19 | -0.10<br>[-3.12, 1.88]<br>p = 0.92 | -0.29 |

**Supplementary Table S2**: Mediation analysis by ipsilesional scalp region using raw thalamus volume (not normative deviation value). Prefrontal, frontal, and central scalp regions showed evidence of partial mediation, although the pre-determined significance threshold of $\alpha = 0.05$ was not met. Model: Independent variable = IL thalamus volume, dependent variable = spectral slowing, mediator = model-estimated Z. Abbreviations: ACME = average causal mediation effect, ADE = average direct effect, Prop. mediated = proportion of total effect via mediation. Square brackets contain 95% confidence intervals. Significance codes: *** p < 0.001 ; ** p < 0.01; * p < 0.05; . p < 0.1
